# Cognitively healthy centenarians are characterized by lower frequencies of six disease-associated *HLA* alleles

**DOI:** 10.1101/2025.03.30.25324899

**Authors:** D. Álvarez Sirvent, N. Tesi, M. Hulsman, A.N. Salazar, N.M. van Schoor, M. Huisman, Y. Pijnenburg, W.M. van der Flier, E.M.M. Strijbis, B.M.J. Uitdehaag, M.J.T. Reinders, S. van der Lee, H. Holstege

## Abstract

**Background:** Human leukocyte antigen (*HLA*) genes are key regulators of immune function and have been implicated in susceptibility to various diseases. However, their role in healthy longevity remains unclear. Here, we investigate the relationship between disease-associated *HLA* alleles and the likelihood of becoming a cognitively healthy centenarian (CHC).

**Methods:** We imputed *HLA* genotypes using genetic data from 3,634 individuals, including 354 CHCs from the Dutch 100-plus Study and 3,269 middle-aged healthy individuals from multiple Dutch cohorts. We examined associations between 59 *HLA* alleles previously linked to 12 diseases—including Alzheimer’s disease (AD), ten autoimmune disorders, and SARS-CoV-2—and the likelihood of becoming a CHC. Logistic regression models were used to estimate odds ratios (ORs), adjusting for population structure. We then calculated centenarian effect ratios (CERs) to compare the effect sizes of *HLA* alleles on the chance of becoming a CHC relative to their known effects on disease susceptibility, assessing the directions of potential pleiotropic effects.

**Results:** While the genomes of CHCs were not enriched with any specific *HLA* alleles, six alleles reduced the likelihood of becoming a CHC (ORs=0.59–0.74, FDR<0.1). These alleles clustered into three haplotypes based on linkage disequilibrium: a class II haplotype (*HLA- DRB1*01:01*, *HLA-DQA1*01:01*, *HLA-DQB1*05:01*), a class I haplotype (*HLA-C*03:04*, *HLA-B*40:01*), and an independent class I allele (*HLA-A*02:01*). We identified both synergistic and antagonistic pleiotropic relationships between autoimmune disease-associated *HLA* alleles and becoming a CHC, with similar magnitudes of effect sizes. *HLA* alleles associated with increased risk of AD consistently exhibited synergistic pleiotropies, with a 5- to 10-fold larger effect on decreased likelihood of healthy longevity.

**Conclusions:** Our findings highlight a complex interplay between *HLA* alleles associated with autoimmune disease susceptibility and healthy longevity. The strongly increased effect sizes and synergistic pleiotropic relationships between the likelihood of becoming a cognitively healthy centenarian and *HLA* alleles previously associated with AD, highlight the involvement of immune-related mechanisms in longevity and neurodegeneration. Further studies, employing Next-Generation-Sequencing and Long-Read-Sequencing, preferably in diverse populations, are needed to map our findings to the full genetic resolution of the *HLA* region.

## Introduction

In the context of an aging global population, the prevalence of age-related diseases is increasing, thereby reducing the quality of life for elderly individuals and posing significant challenges to healthcare systems worldwide (1). A key factor in aging is the decline in the responsiveness of the immune system, suggesting that an impaired immune system may accelerate the aging process itself (2). Conversely, preserved immune system function could promote longevity and overall health, which is supported by the observation that human longevity is linked to a lower genetic risk for age-related diseases involving the immune system (3,4).

The human leukocyte antigen (*HLA*) region, located in chromosome 6, is critical for immunity and stands as the most polymorphic region in the human genome (5). Genetic polymorphisms in *HLA* have been associated with autoimmune diseases (6–8) as well as cancer (9), cardiovascular diseases (10), metabolic and digestive diseases, and neurodegenerative diseases (11), such as Alzheimer’s disease (AD) (12,13) and Multiple Sclerosis (MS) (14). This makes the *HLA* region the one with the most disease associations based on Genome-Wide Association Studies (GWAS), highlighting the potential impact of immune functioning on disease susceptibility and human longevity.

Of all age-related diseases, AD is one of the most prevalent, representing the most common form of dementia at old age (15). Despite greater odds of developing AD at old age (16), a rare subset of individuals demonstrates exceptional longevity with sustained cognitive function, providing insights into genetic mechanisms underlying this phenomenon (17). Recently, we showed that cognitively healthy centenarians (CHCs), i.e. those older than 100 years who self-report to be cognitively healthy, are genetically protected against AD (18).

Interestingly, several protective Single Nucleotide Polymorphisms (SNPs) were more strongly enriched in CHCs than others, with many functionally linked to the immune system. This suggests an important role of immune-related processes in the chance of becoming a CHC. One of the protective SNP that was among the strongest enriched in the CHCs was in the *HLA* region further corroborating with the involvement of the immune system (12).

The *HLA* region consists of seven so-called classical *HLA* genes, divided into three class I genes (*HLA-A*, *HLA-B*, *HLA-C*) and four class II genes (*HLA-DRB1*, *HLA-DQA1*, *HLA-DQB1*, *HLA-DPB1*). These genes encode histocompatibility molecules that are essential for the immune system’s recognition of self and foreign substances (19,20). By presenting antigenic peptides to T-cells, they play a central role in both adaptive and innate immune responses, influencing infection, autoimmunity, and overall immune regulation (21). *HLA* alleles, genetic variants or versions of *HLA* genes, have unique names. Each allele name starts with the *HLA* prefix, a hyphen, the gene name and “*” as a separator (for example “*HLA-A\**”), followed by up to four ‘fields’ of digits separated by colons, each successive field representing a deeper level of genetic resolution in the allele sequence (22). The first two fields correspond to respectively the serological type and the *HLA*-protein sequence, for example “*HLA-A*02:01*". SNP-based GWAS have been crucial for identifying human trait associations in *HLA* loci. However, the extreme polymorphism and complex Linkage Disequilibrium (LD) patterns in this region complicate SNP-only analyses, as SNPs alone cannot fully represent this diversity. Additionally, *HLA* SNPs often deviate from Hardy-Weinberg equilibrium, leading to their exclusion from genotyping arrays (23). In contrast, *HLA* alleles at a two-fields resolution define distinct protein sequences that offer greater biological insight than SNPs, enhancing our understanding of human population diversity. To better explore associations between *HLA* polymorphisms and human traits, it is essential to analyze *HLA* alleles at the highest resolution available while considering LD. Here, we employed *HLA* imputation with population-based reference models to infer *HLA* alleles at up to two-fields resolution (24).

Previous studies have suggested that carrying specific *HLA* alleles may contribute to extreme longevity. The *HLA-DRB1*15* allele was enriched in Sardinian centenarians (25), and an intergenic SNP located between *HLA-DRB1* and *HLA-DQA1* was associated with longevity (26). More recently, specific subtypes of *HLA-DRB4*04* that were linked to an *HLA* SNP enriched in the CHCs were identified to have protective effects against AD (13). Furthermore, a common *HLA-B* allele was shown to be associated with an increased chance of asymptomatic SARS-CoV-2 infection (27). Meanwhile, eight other *HLA* alleles were significantly associated with increased risk of AD in the latest AD GWAS (12), and a recent UK Biobank (UKB) study identified 129 novel associations between *HLA* alleles and autoimmune diseases (6).

Despite these associations with disease, a gap remains in our understanding of how disease-associated *HLA* alleles influence longevity, particularly in individuals who maintain cognitive health. Here, we set out to compare the frequencies of previously disease-associated *HLA* alleles in Dutch CHCs with those in middle-aged healthy individuals. Additionally, we explored pleiotropic relationships by investigating whether *HLA* alleles associated with AD, several autoimmune diseases, and SARS-CoV-2 infection, also affect the chance to become a CHC.

## Methods

### Study participants

Genetic data from a total of 4,151 individuals was included in this study. Of these, 379 were self-reported CHCs from the 100-plus Study cohort (17). These are Dutch individuals aged 100 years or older, who self-reported to be cognitively healthy. We compared CHCs to 3,772 middle-aged individuals healthy at blood draw, from different study cohorts: 1,773 community dwelling older adults from Longitudinal Aging Study of Amsterdam (LASA) (28), 1,524 individuals from Amsterdam Dementia Cohort (ADC), who were labeled cognitively healthy after extensive neurological examination (29), 62 healthy controls from the Netherlands Brain Bank (30), 95 healthy individuals (partners of CHCs’ offspring) from the 100-plus Study, 195 healthy individuals from the Netherlands Twin study (31), and 123 healthy individuals from the Project Y cohort (32). The Medical Ethics Committee of the Amsterdam UMC approved all studies. All participants and/or their legal representatives provided written informed consent for participation in clinical and genetic studies.

### Genotyping, SNP imputation and Quality Control (QC)

Genetic variants were determined by standard genotyping and imputation methods, and applying established QC methods (33). We used high-quality genotypes in all individuals (individual call rate >99%, variant call rate >99%). Departure from Hardy-Weinberg equilibrium was considered significant at p<1x10-6. We excluded individuals with sex mismatches. Before downstream analyses, we excluded individuals with a family relation (identity-by-descent >= 0.2) (34), and we excluded individuals of non-European ancestry (based on 1000Genomes clustering (35), leaving 3623 individuals in the analysis.

### Model-based HLA imputation

HLA genotypes at two-fields allele resolution were imputed using the software HIBAG (36) and the prediction model specific to our population (of European ancestry) and genotyping platform, *Illumina Infinium Global Screen Array version 2.0.* This model imputes alleles for all seven classical *HLA* class I and class II genes. Imputation was performed for all individuals retained after QC. After imputation, only genotypes with posterior probability >0.5 were kept, as recommended by authors.

### Inclusion of allele-disease associations

We focused on the associations between *HLA* alleles and diseases that were previously reported from three research articles (6,12,27) (Figure 1.A). These included 11 autoimmune diseases from the UK Biobank (UKB) study, AD from the latest GWAS, and SARS-CoV-2 from a study of a National Marrow Donor Program cohort (United States). From the UKB study, effect sizes and standard deviations of associations (at two-fields resolution, based on European ancestry) were extracted from Supplementary Data 20 (full summary statistics), with a significance threshold as defined in the paper (p-value < 5e-8/11) (6). For AD, summary statistics for all *HLA* allele associations reported in Supplementary Table 8 were extracted (12). For SARS-CoV-2, association statistics from the allele that was significantly associated with an asymptomatic infection were used (27). Alleles with frequency lower than 0.01 (in original paper and/or in our study cohort) and alleles that could not be called by applied imputation models were excluded.

**Figure 1.**
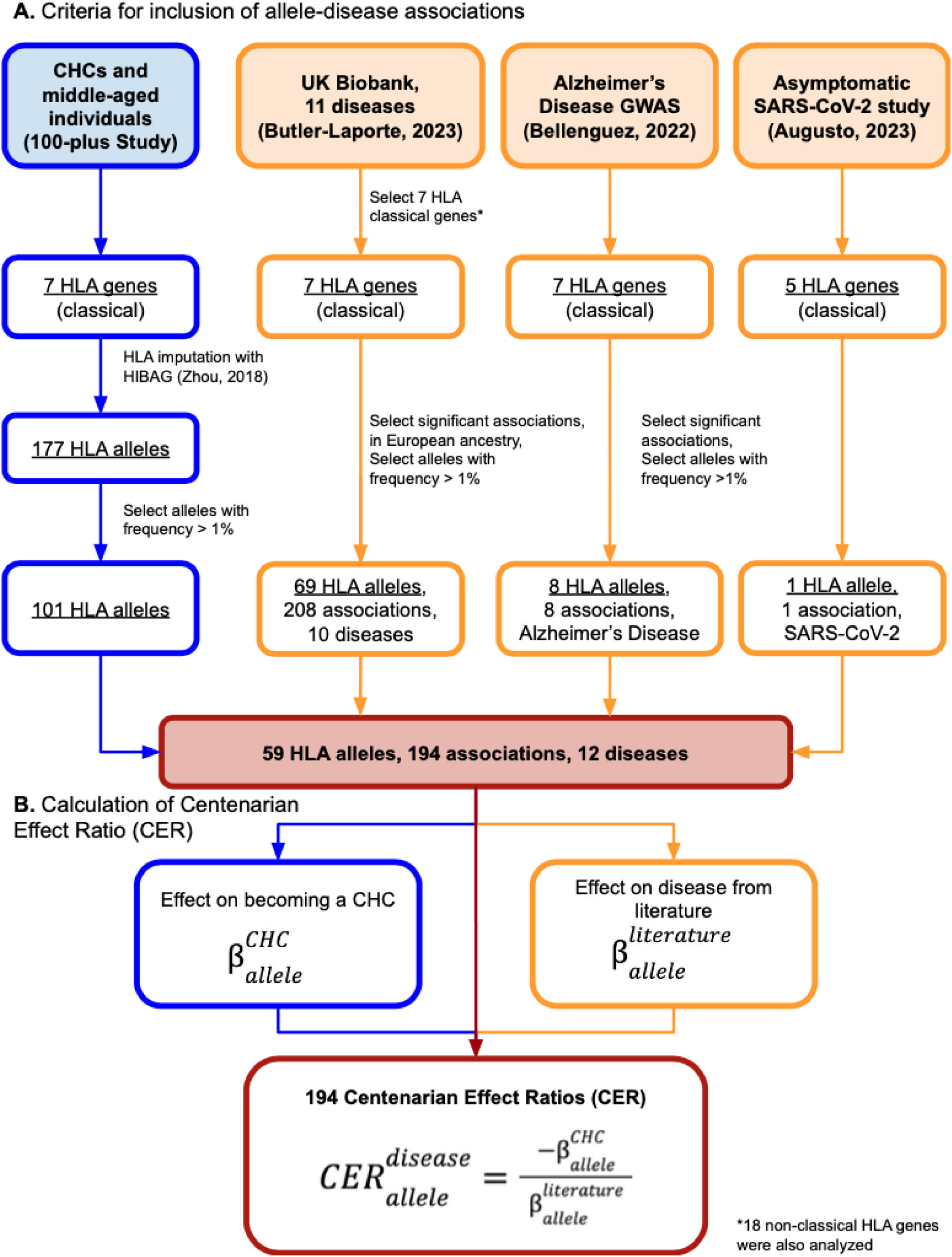
Study workflow. A) criteria for inclusion of allele-disease associations, B) calculation of Centenarian Effect Ratio (CER).

In total, 59 unique alleles were included (Figure 1.A): eight associations for AD, one for SARS-CoV-2, and 185 for ten out of the 11 autoimmune diseases, summing to 194 associations with 12 diseases. Of the autoimmune diseases, 37 *HLA* alleles associated with autoimmune thyroid disorders (ATD), 31 to Celiac disease (CD), 26 to Psoriasis, 22 to Asthma, 20 to Rheumatoid Arthritis (RA), 17 to Type 1 Diabetes (T1D), 12 to Polymyalgia Rheumatica or Giant Cell Arteritis (PMR-GCA), nine to Ankylosing Spondylitis (AS), nine to MS and demyelinating disease (MS), and two to Ulcerative Colitis (UC).

### Association between HLA alleles and becoming a CHC

We studied the frequencies of disease-associated *HLA* alleles in CHCs compared to middle-aged healthy individuals, to obtain the effect of an allele on becoming a CHC: 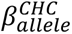 (or 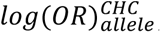. Effects were estimated using logistic regression, with CHC as “cases” and middle-aged healthy individuals as “controls”. Regression models were corrected for population effect (first five genetic principal components) (37,38), and p-values of associations were FDR corrected for multiple testing (39).

### Centenarian effect ratio (CER) calculation

To compare the effect on becoming a CHC with the reported effect on disease, we calculated the centenarian effect ratio (CER, Figure 1.B) per *HLA* allele-disease association (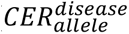). To calculate this, we took the effect (beta of regressor) of *not* becoming a CHC of each allele (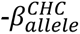), and divided it by the published effect of the allele on the disease (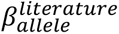), as reported in the reference manuscripts.

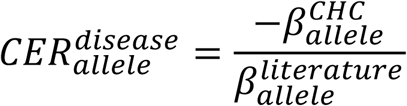

A positive CER (CER>0) indicates that carrying a given *HLA* allele is associated with a decreased chance of becoming a CHC and simultaneously with an increased chance of developing a disease, or vice-versa (i.e. increased chance of becoming a CHC and decreased chance of developing disease). Therefore, an allele with a positive CER shows a synergistic pleiotropy, i.e. affects positively or negatively both healthy longevity and avoiding disease. A negative CER (CER<0) indicates that carrying the *HLA* allele is associated with a decreased chance of becoming a CHC but at the same time with a decreased chance of developing a disease, or vice-versa (i.e. increased chance for becoming a CHC but increased chance for a disease). An HLA allele with a negative CER shows an antagonistic pleiotropy, i.e. increasing (or decreasing) healthy longevity but at the same time also increasing (or decreasing) disease risk. Confidence intervals of the CER, and the probability of divergence of the CER from a value of 0 (i.e. either synergistic or antagonistic pleiotropy) were estimated using sampling techniques (S=10,000, with a two-tailed p-value). Two-tailed p-values were corrected for multiple testing using the FDR method.

### Linkage Disequilibrium calculation

Linkage Disequilibrium (LD) scores (quantified with *r*^2^) were calculated for all pairwise combinations of *HLA* alleles from different genes, using the software PLINK (40). All statistical analyses were performed using custom R Statistical Software scripts, unless stated otherwise.

## Results

### Six HLA alleles were associated with decreased odds of becoming a cognitively healthy centenarian (CHC)

After genotyping, *HLA* allele imputation (posterior probability >0.5) and QC, our study cohort consisted of N=354 CHCs (age 101.05 ± 2.51, 71% females) and N=3,269 middle-aged healthy individuals (age 62.57 ± 8.66, 48% females). Of all 25,361 imputed *HLA* genotypes across seven *HLA* genes, 97.5% were accepted for downstream analysis.

For the 59 unique *HLA* alleles that were included in the 194 unique allele-disease associations, we calculated the effect sizes associated with becoming a CHC relative to middle-aged healthy individuals. We did not identify *HLA* alleles that were more frequent in CHCs. However, we identified six unique alleles, with frequencies ranging from 0.07 to 0.3 in our cohort groups, that were significantly less frequent in CHCs. These six alleles were associated with 41% to 26% decreased odds of becoming a CHC (FDR<0.1): i.e. odds ratios (ORs) ranging from 0.59-0.74 (Table 1, Figure 2).

**Figure 2.**
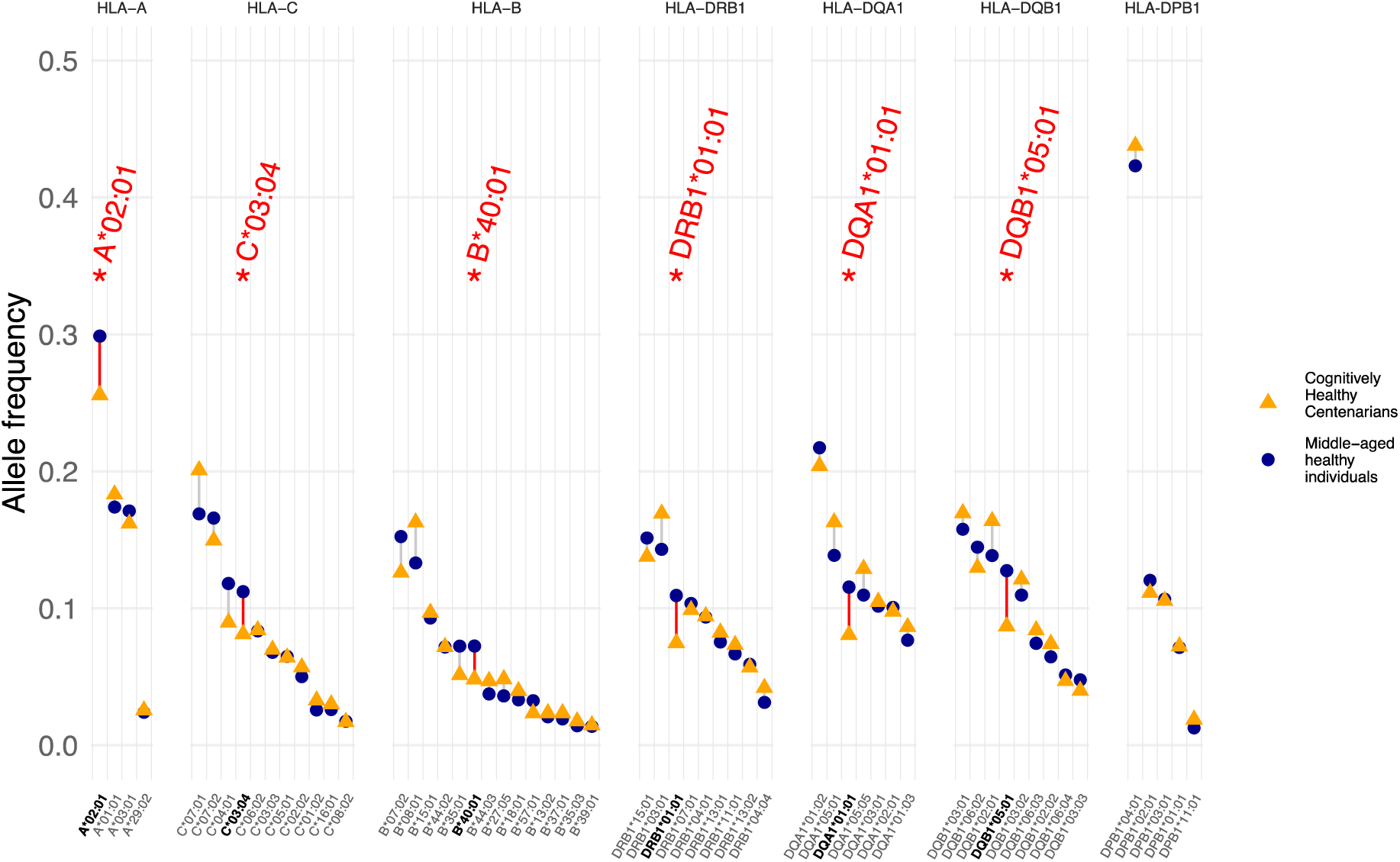
Allele frequencies of 59 tested alleles in cognitively healthy centenarians (orange triangles) and middle-aged healthy individuals (blue circles). The six alleles that were significantly less frequent in CHCs (FDR<0.1) have bold labels and are highlighted in red.

**Table 1.** Summary statistics for the six alleles significantly associated with decreased odds of becoming a CHC. Allele name (Allele), Odds Ratio of becoming a CHC with 95% confidence intervals (ORs CHCs [CIs]), allele frequency in CHC (Freq. CHCs), allele frequency in middle-aged healthy individuals (Freq. Controls), p-value of association (p-value), false-discovery-rate (FDR) corrected p-value of association, disease, Centenarian Effect Ratio with 95% CI (CER [CIs]), OR of disease with 95% CI (ORs disease [CIs]).

| Allele | ORs CHCs [CIs] | Freq. CHCs | Freq. Controls | p-value | FDR | Disease | CER [CIs] | ORs disease [CIs] |
| --- | --- | --- | --- | --- | --- | --- | --- | --- |
| *HLA-A\*02:01* | 0.74 [0.6-0.91] | 0.26 | 0.3 | 0.004 | 0.072 | AD | 10.36 [3.18 - 31.8] | 1.03 [1.01 - 1.05] |
|  |  |  |  |  |  | Psoriasis | 1.7 [0.52 - 2.98] | 1.2 [1.15 - 1.24] |
|  |  |  |  |  |  | CD | 1.38 [0.43 - 2.64] | 1.25 [1.16 - 1.33] |
|  |  |  |  |  |  | MS | -0.9 [-1.6 - -0.28] | 0.71 [0.66 - 0.76] |
| *HLA-C\*03:04* | 0.6 [0.43-0.83] | 0.08 | 0.11 | 0.002 | 0.072 | PMR-GCA | 1.68 [0.59 - 2.93] | 1.36 [1.25 - 1.47] |
|  |  |  |  |  |  | T1D | 1.39 [0.51 - 2.43] | 1.45 [1.33 - 1.57] |
|  |  |  |  |  |  | Psoriasis | -2.44 [-4.51 - -0.84] | 0.81 [0.76 - 0.86] |
| *HLA-B\*40:01* | 0.59 [0.4-0.87] | 0.05 | 0.07 | 0.007 | 0.089 | PMR-GCA | 1.59 [0.43 - 3] | 1.4 [1.27 - 1.52] |
| *HLA-DRB1\*01:01* | 0.63 [0.44-0.89] | 0.07 | 0.11 | 0.001 | 0.097 | AD | 5.36 [1.28 - 11.82] | 1.09 [1.05 - 1.13] |
|  |  |  |  |  |  | AS | 1.32 [0.32 - 2.64] | 1.42 [1.26 - 1.58] |
|  |  |  |  |  |  | CD | -1.2 [-2.35 - -0.29] | 0.68 [0.6 - 0.76] |
|  |  |  |  |  |  | ATD | -3.08 [-5.66 - -0.73] | 0.86 [0.83 - 0.89] |
|  |  |  |  |  |  | Asthma | -4.56 [-8.61 - -1.11] | 0.9 [0.88 - 0.92] |
| *HLA-DQA1\*01:01* | 0.64 [0.46-0.89] | 0.08 | 0.12 | 0.008 | 0.089 | AD | 7.67 [2.01 - 17.8] | 1.06 [1.03 - 1.09] |
|  |  |  |  |  |  | AS | 1.39 [0.38 - 2.72] | 1.38 [1.25 - 1.51] |
|  |  |  |  |  |  | CD | -0.91 [-1.66 - -0.24] | 0.61 [0.55 - 0.67] |
|  |  |  |  |  |  | Asthma | -4.88 [-8.9 - -1.27] | 0.91 [0.9 - 0.93] |
|  |  |  |  |  |  | ATD | -5.97 [-12.07 - -1.55] | 0.93 [0.9 - 0.95] |
| *HLA-DQB1\*05:01* | 0.63 [0.46-0.86] | 0.09 | 0.13 | 0.003 | 0.072 | AD | 8.02 [2.49 - 18.6] | 1.06 [1.03 - 1.09] |
|  |  |  |  |  |  | CD | -1.02 [-1.91 - -0.33] | 0.63 [0.55 - 0.71] |
|  |  |  |  |  |  | Asthma | -5.35 [-9.98 - -1.69] | 0.92 [0.89 - 0.94] |

Based on Linkage Disequilibrium (LD) in our cohort, these six alleles could be clustered in three groups (Figure 3): a class II haplotype (combination of three alleles) encompassing *HLA- DRB1*01:01*, *HLA-DQA1*01:01* and *HLA-DQB1*05:01*, in high LD (*r*^2^ values from 0.8 to 0.91); a class I haplotype (combination of two alleles) encompassing *HLA-C*03:04* and *HLA-B*40:01*, in moderate LD (*r*^2^=0.58); and a class I allele *HLA-A*02:01*, which is not in LD with any of the other significant alleles. The high LD in the class II haplotype indicates that any of the three alleles could be driving the observed effect on becoming a CHC, with no significant differences between their effects.

**Figure 3.**
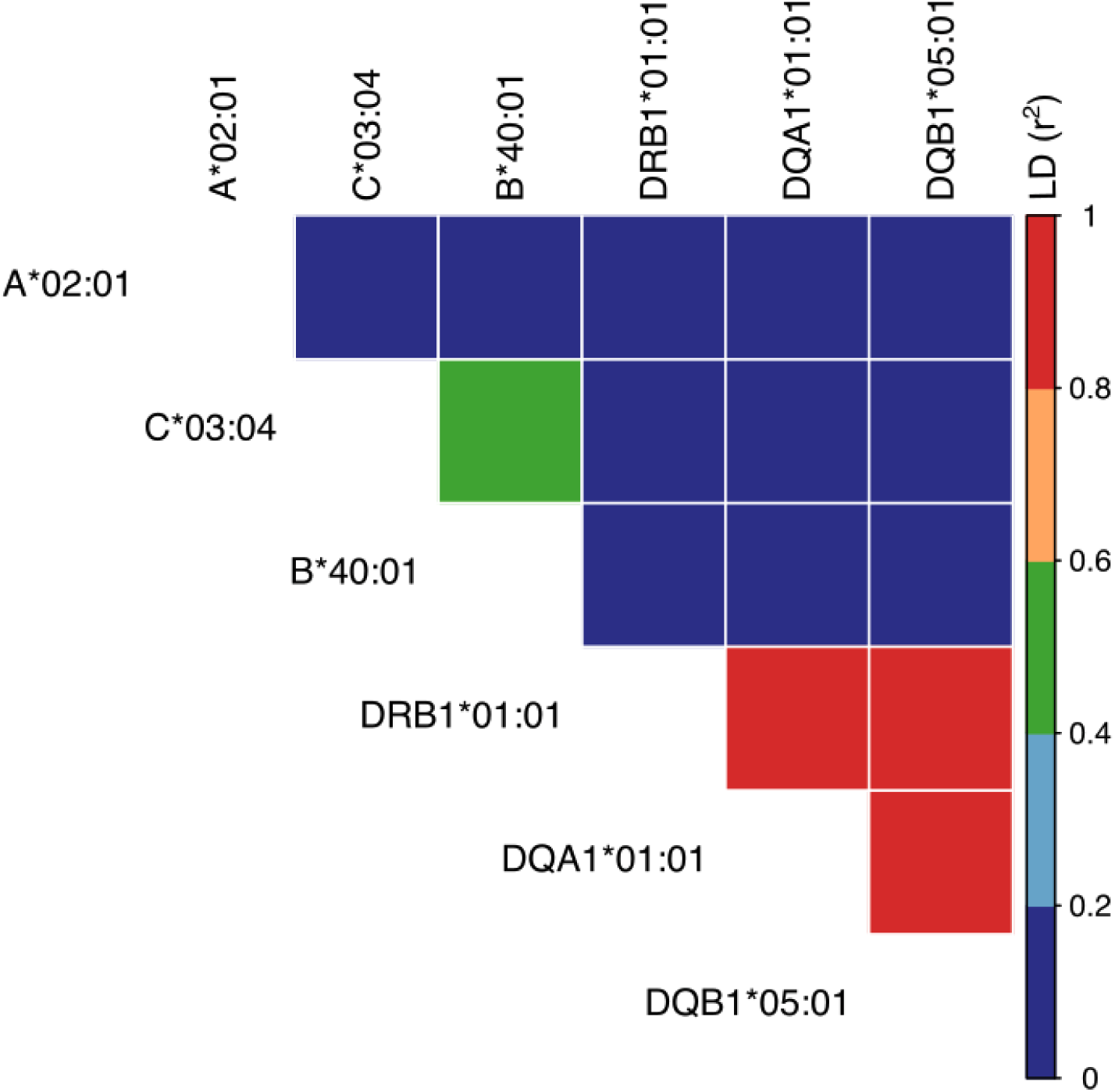
Linkage Disequilibrium (r^2^) matrix of the six alleles significantly associated with decreased odds of becoming a CHC.

The *HLA-C*03:04* and *HLA-B*40:01* alleles in the class I haplotype are in moderate LD, and their effect sizes for becoming a CHC were very similar (ORs=0.59 and 0.6). However, examining allele frequencies in our study cohort revealed that 98% of the carriers of *HLA- B*40:01* (frequency=0.07) also carried *HLA-C*03:04* (frequency=0.10), while only 70% of the *HLA-C*03:04* carriers carried the *HLA-B*40:01* allele. This suggests that *HLA-C*03:04* may be the primary driver of the observed effect on becoming a CHC, while the effect of *HLA- B*40:01* could be a passenger-effect attributable to its LD with *HLA-C*03:04*. Moreover, *HLA- C*03:04* is not in LD with any other alleles at r² > 0.1 (including other *HLA-B* alleles), making it unlikely that the observed effect is due to alleles with lower LD.

### The six HLA alleles associated with decreased odds of becoming a CHC had 21 associations in nine diseases, with significant pleiotropies

For each of the 194 allele-disease associations (59 *HLA* alleles and 12 diseases), we assessed pleiotropic relationships between the known effects of the alleles on disease, and their effects on becoming a CHC. For this, we calculated Centenarian Effect Ratios (CERs) by dividing the effects on *not* becoming a CHC by the known effects on disease as obtained from literature. We first focused on the CERs of the six alleles that were significantly less frequent in CHCs compared to middle-aged healthy individuals. These were involved in 21 significant allele-disease associations, covering nine diseases, in which CERs diverged significantly from 0 (Figure 4). Of these associations, 11 had a positive CER, while ten had a negative CER (Figure 4, Table 1). All alleles, except *HLA-B*40:01*, had both positive and negative CERs, meaning that they were involved both in synergistic and antagonistic pleiotropic effects on becoming a CHC and disease susceptibility.

**Figure 4.**
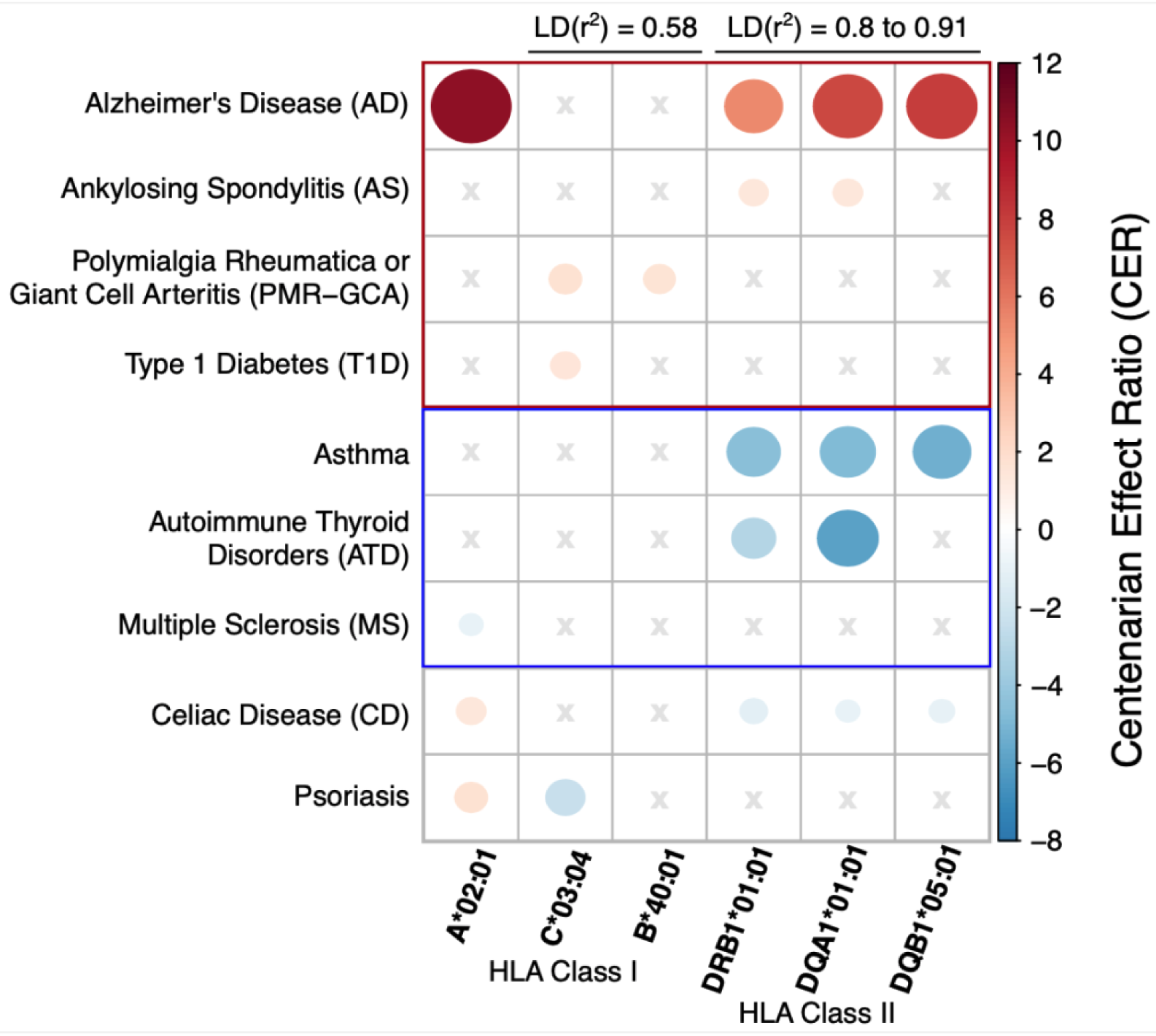
Centenarian Effect Ratios (CERs) for disease associations of the six alleles significantly associated with decreased odds of becoming a CHC. The size of the dots indicates the magnitude of the effect observed in the centenarians, compared to the effects reported in literature. Red dots indicate relationships of synergistic pleiotropy: HLA alleles increasing disease risk and decreasing the chance to become a CHC, and the red rectangle marks diseases that showed exclusively synergistic pleiotropies. Blue dots indicate antagonistic pleiotropy: HLA alleles decreasing disease risk and decreasing the chances of becoming a CHC, and the blue rectangle marks diseases that showed exclusively antagonistic pleiotropies. Disease types with no significant associations (SARS-CoV-2, UC, RA) are not shown. Grey crosses indicate no prior significant association between allele and disease.

We observed exclusively significantly positive CERs (CER>0, red rectangle in figure 4), i.e. synergistic pleiotropies, among (i) associations with AD, in class II haplotype alleles *HLA- DRB1*01:01*, *HLA-DQA1*01:01* and *HLA-DQB1*05:01*, and class I allele *HLA-A*02:01* (CERs ranging from 5.4 to 10.4), (ii) associations with PMR-GCA in class I haplotype alleles *HLA- C*03:04* and *HLA-B*40:01* (CERs=1.6 & 1.7), (iii) associations with AS in class II haplotype alleles *HLA-DRB1*01:01* and *HLA-DQA1*01:01* (CERs=1.32 & 1.39,) and (iv) an association with T1D in class I haplotype allele *HLA-C*03:04* (CER=1.39). These alleles were associated with decreased likelihood of developing these diseases, and with a decreased chance of becoming a CHC. Notably, for the alleles associated with AD, the effect size of the association with a decreased risk of becoming a CHC was substantially larger than the effect size associated with increased AD risk, i.e. five to ten-fold larger.

Interestingly, for all alleles except *HLA-B*40:01* (which was only associated with increased risk of PMR-GCA), we also observed significantly negative CERs (CER<0), i.e. antagonistic pleiotropies. These were exclusively observed (figure 4, blue rectangle) with diseases such as (i) asthma in class II haplotype alleles (CERs=-5.4 to -4.6), (ii) ATD in class II haplotype alleles *HLA-DRB1*01:01* and *HLA-DQA1*01:01* (CERs=-6 & -3.1) and (iii) MS in *HLA-A*02:01* (CER=-0.9). This implies that despite having protective effects against diseases, these alleles were also associated with *decreased* chances of becoming a CHC.

Alleles associated with CD and Psoriasis had significantly positive as well as negative CERs, i.e. synergistic and antagonistic pleiotropies: CD with four alleles (negative CERs from -1.2 to -0.91 for *HLA-DRB1*01:01*, *HLA-DQA1*01:01* and *HLA-DQB1*05:01*, and positive CER=1.38 for *HLA-A*02:01*) and Psoriasis with two (negative CER=-2.44 for C*03:04, positive CER=1.7 for *HLA-A*02:01*). Alleles associated with RA, UC and SARS-CoV-2 were not included in the six alleles that were significantly less frequent in CHC (and are thus excluded from Figure 4), nor had significant CERs -i.e. no pleiotropic relationships with becoming a CHC (Figure 5).

**Figure 5.**
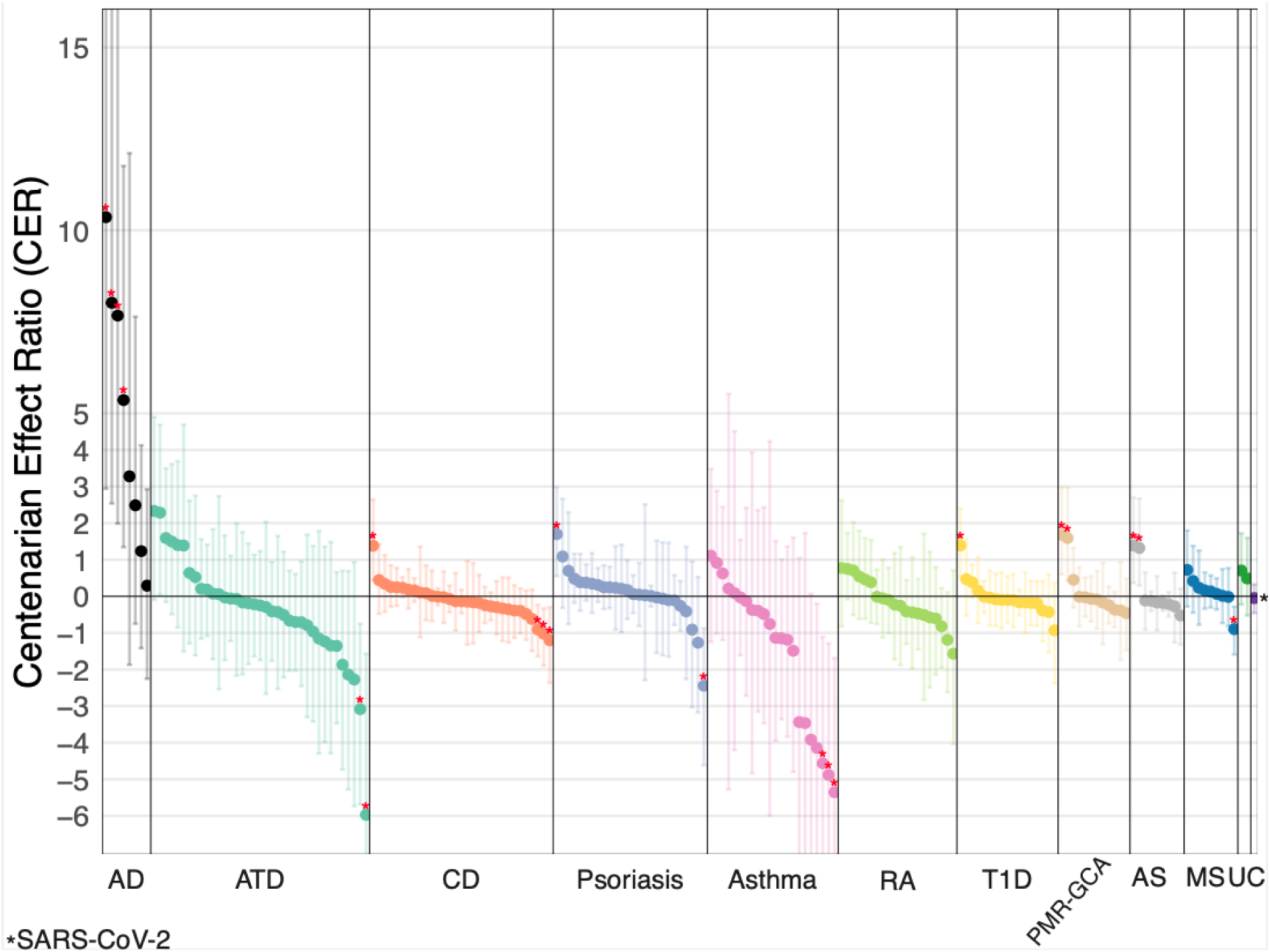
Centenarian Effect Ratios (CERs) with 95% confidence intervals (CIs) for all 194 allele-disease associations. Positive CERs indicate synergistic pleiotropy relationships between disease associations and becoming a CHC, negative CERs indicate antagonistic pleiotropy. Red stars mark values significantly deviated from 0 (FDR<0.1). 12 diseases, differentially colored from left to right: Alzheimer’s Disease (AD), autoimmune thyroid disorders (ATD), Celiac disease (CD), Psoriasis, Asthma, Rheumatoid Arthritis (RA), Type 1 Diabetes (T1D), Polymyalgia Rheumatica or Giant Cell Arteritis (PMR-GCA), Ankylosing Spondylitis (AS), Multiple Sclerosis and demyelinating disease (MS), Ulcerative Colitis (UC) and SARS- CoV-2. Allele names and supporting information is available in Additional Table 1.

### Associations with AD had consistently positive and larger Centenarian Effect Ratios (CERs) than all other diseases

To contextualize the pleiotropic associations of the six alleles that were significantly less frequent in CHCs compared to middle-aged individuals, we examined the CER values for all 194 allele–disease associations across 12 diseases (Figure 5). The CERs for most allele-disease associations were close to 0, meaning that they did not show consistent pleiotropic relationships between becoming a CHC and getting a disease. In contrast, all eight *HLA* alleles linked to AD had positive CERs, showing a consistent synergistic pleiotropy. The median CER for AD associations was 4.4, notably higher than the medians for other diseases (between −0.9 and 0.6), underscoring that the effect of AD risk on the chance of becoming a cognitively healthy centenarian is substantial.

Four AD risk-increasing alleles were significantly less frequent in CHCs (*HLA-A*02:01*, *HLA- DRB1*01:01*, *HLA-DQA1*01:01* and *HLA-DQB1*05:01*) with no major pleiotropic effects (Figure 6A). The one AD risk-increasing allele was not significantly less frequent in CHCs (class I allele *HLA-B*57:01*), and the three AD-protective alleles, class II alleles *HLA- DRB1*04:04*, *HLA-DQA1*03:01* and *HLA-DQB1*03:02* (in high LD, r^2^=0.9 in our cohort), were more frequent in CHCs, albeit not significant. The lack of significance of these four alleles may be due to lower allele frequencies in our cohort (0.031–0.042, Additional Table 1), than the frequencies of the found significant alleles (0.07–0.3, Table 1). However, the substantially lower CERs associated with these alleles may be explained by major pleiotropic effects in other diseases: while class II alleles *HLA-DRB1*04:04* and *HLA-DQB1*03:02* are both associated with small protective effects against AD (OR = 0.88 and 0.90), they are also the major risk-increasing alleles for PMR-GCA (*HLA-DRB1*04:01*, OR = 2.4) and T1D (*HLA- DQB1*03:02*, OR = 2.2) (Figure 6B). Furthermore, while *HLA-B*57:01* was associated with a limited risk-increasing effect on AD (OR = 1.1), it has a strong risk-increasing effect for psoriasis (OR = 3.0) in combination with a strong protective effect against CD which may have contributed to becoming a CHC (OR = 0.42) (Figure 6B). We speculate that similar interactions may explain why alleles with the strongest disease associations did not have significantly increased or decreased frequencies in CHCs (Additional figure 1).

**Figure 6.**
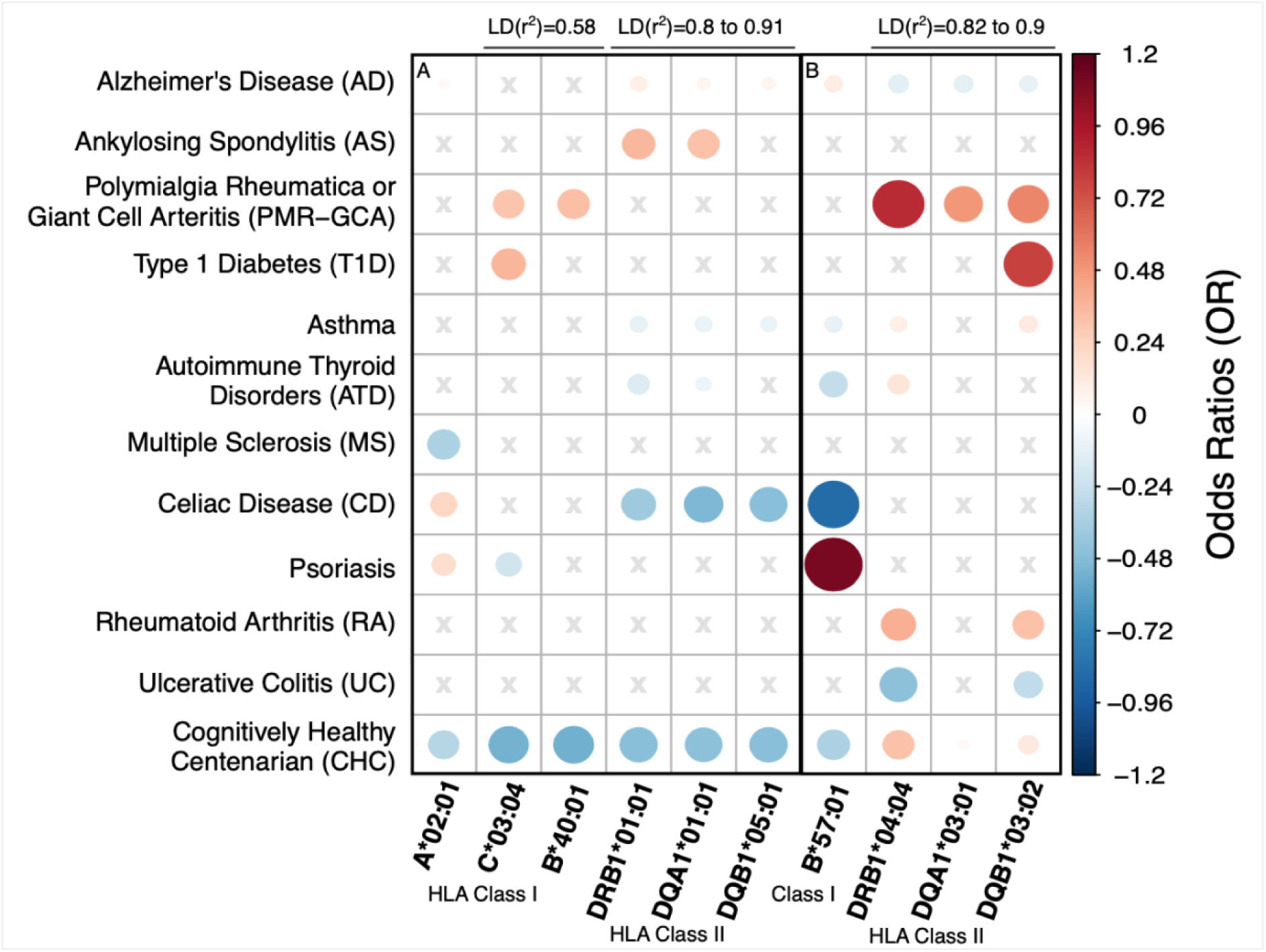
Odds Ratios (ORs) for literature-reported associated diseases (first 11 columns) and becoming a CHC (CHC column), per allele. **A)** ORs of the six alleles significantly associated with decreased odds of becoming a CHC. **B)** ORs of four alleles associated with AD in literature, which were not significantly associated with becoming a CHC. The size of the dots indicates the magnitude of the effect size: red dots indicate odds-increasing associations whereas blue dots indicate the odds-decreasing associations. Disease types with no significant associations (SARS-CoV-2) are not shown. Grey crosses indicate no prior significant association between allele and disease.

## Discussion

In this study, we found that six *HLA* alleles, that were previously associated with Alzheimer’s Disease and eight autoimmune diseases, were also significantly associated with a lower likelihood of reaching extreme ages while maintaining cognitive health. The effect sizes of these alleles of having a lower chance to become a cognitively healthy centenarian (CHC) were either in the same or opposite direction as their effect on risk to develop autoimmune disease, and were similar in magnitude. This suggests that there is not a direct relationship between increased risk of these diseases and lower chance of becoming a CHC. However, for four *HLA* alleles associated with increased risk of developing AD, the effects on becoming a CHC were far stronger than the effects on the increased risk of getting AD: class I allele *HLA-A*02:01* and a haplotype encompassing three *HLA* class II alleles (*HLA-DRB1*01:01*, *HLA-DQA1*01:01* and *HLA-DQB1*05:01*). This suggests that these *HLA* alleles have a strongly unfavorable functional effect that goes beyond AD-related mechanisms and influences general survival mechanisms.

Our observation that *HLA* alleles associated with increased AD risk were depleted in CHCs was according to expectations since CHCs were selected for their cognitive health despite their extreme age. However, the strong five to ten fold increase in effect size of the association with becoming a CHC compared to the effect size of the association with AD (18) is extraordinary. Although, it fits with our previous observations that genomes of healthy centenarians are maximally enriched with protective variants against AD and depleted with risk-increasing variants (41). This may be explained by pleiotropic effects of these alleles, i.e. these alleles also show associations with other seemingly independent disease phenotypes. For example, we previously observed that the rs72824905-G variant in the immune-linked *PLCG2* gene, associated with protection against AD (43), was also associated with protection against other forms of dementias, including Dementia with Lewy Bodies, Frontotemporal Dementia (44), with MS (45), and strongly with becoming a CHC.

Indeed, for the six *HLA* alleles that were significantly less frequent in CHCs compared to middle-aged healthy individuals, we observed both synergistic and antagonistic pleiotropies with respect to observed associations with eight autoimmune diseases (6). Upon inspection of the pleiotropic associations that might drive the effects on a lower chance to become a CHC, we observed that *HLA* alleles associated with increased risk of AD and Polymyalgia Rheumatica or Giant Cell Arteritis (PMR-GCA) had exclusively synergistic pleiotropies. Intriguingly, these diseases generally develop after 50 years (15,42), suggesting that an immune system with a higher genetic risk of developing age-related diseases may contribute to a decreased chance of healthy longevity. On the other hand, diseases for which we observe exclusively antagonistic pleiotropic effects, including asthma (43), autoimmune thyroid disorders (ATD) (44) and Multiple Sclerosis (MS) (45) generally develop before 40 years. One can speculate that an immune system with an increased genetic risk for autoimmune disease at a younger age, may have advantageous effects at old ages, where a more active immune system may contribute to maintain health, while immunogenetic protection against these conditions may lead to a decreased chance of becoming a CHC. This is in analogy to higher blood pressure being a risk factor for AD at younger age, but protective against AD at older age (46). Clearly, this concept requires further research, as we observe exceptions: while the onset of Type 1 Diabetes (T1D) and Ankylosing Spondylitis occurs generally before 30 years (47,48), we observed exclusively synergistic pleiotropies among the *HLA* alleles that significantly associated with becoming a CHC, suggesting that an *HLA*-related higher risk of these early onset diseases leads to a life-long decreased chance of becoming a CHC. On the other hand, none of the six alleles that were significantly associated with decreased chances of becoming a CHC, were associated with Rheumatoid Arthritis (RA), despite symptoms developing at later age and its clear immunogenetic links (7), suggesting that *HLA*-related development of RA does not influence the chances of longevity. Furthermore, we observed both synergistic and antagonistic pleiotropic effects in five of the significant alleles with psoriasis and Celiac Disease, which can develop throughout life (49,50). Together, the diversity of these pleiotropic relationships complicates the interpretation of the contributing effects of *HLA* disease-associations on becoming a CHC in our cohort.

The remaining disease-associated *HLA* alleles were not significantly associated with becoming a CHC. Even the *HLA* alleles with the strongest effects on the susceptibility of autoimmune diseases (e.g. *HLA-DRB1*15:01* for MS (14) *HLA-B*27:05* for AS (8)) and SARS-CoV-2 did not overlap with the alleles that were significantly associated with becoming a CHC. This implies that CHCs were neither consistently at higher nor lower risk for those diseases based on *HLA* genetics, suggesting that protection against these diseases, at least in the *HLA* region, does not play a major role on healthy longevity. We specifically considered the AD-associated class II *HLA-DRB1*04:04* allele, which was previously suggested to drive the protective effect of the class II haplotype with *HLA-DRB1*04:04*, *HLA-DQA1*03:01* and *HLA- DQB1*03:02* against AD (13), but we observe only a non-significant enrichment of this haplotype in CHCs. Possibly, the allele frequency of *HLA-DRB1*04:04* was too low to observe a significant effect in our cohort. In addition, the protective effect of this haplotype may be more prominent when assessed in the context of AD only, while in context of longevity the effect may be compromised by its strong association with increased risk for developing PMR-GCA and T1D, and possibly other diseases. Furthermore, the SARS-CoV-2 association stands out: since almost all centenarians survived COVID-19 without severe symptoms, it was in contrast to our expectations that they were not enriched with the *HLA-B*15:01* allele, which was associated with a 2.4 fold increased chance of asymptomatic SARS-CoV-2 infection (27). Apparently, the resilience to COVID-19 in these centenarians may have involved other, yet unidentified factors. Future studies will have to assess if the net effect of this allele on longevity has changed after the pandemic.

The limited sample size of cognitively healthy centenarians impelled us to maximize the statistical power of our analysis by focusing on relatively common (>1% frequency) *HLA* alleles that were previously associated with diseases. We could not provide a full review of HLA alleles associated with healthy human longevity, as we could not calculate CERs for unknown associations. However, we cannot exclude that there are undiscovered disease associations that might show more pleiotropies with becoming a CHC. Furthermore, we warn that our Centenarian Effect Ratio (CER) values may be slightly inflated. They depend on effect sizes determined by large sample size studies, which, due to the inclusion of by-proxy cases, may be diluted, relative to when focusing only on clinical cases. While *HLA* imputation from SNP genotyping data allows genotyping at two-fields *HLA* allele resolution (i.e. protein sequence level) in classical *HLA* genes, future studies are warranted to further fine-map our findings to the full genomic resolution of the *HLA* region, for example by using long-read sequencing techniques. Lastly, the individuals in this study have a European/Dutch population background. Given the large variability in *HLA* alleles across different ancestries, our findings may not generalize to populations with different *HLA* architectures.

## Conclusions

We identified six *HLA* alleles that were significantly less frequent in cognitively healthy centenarians (CHCs) compared to middle-aged healthy individuals, suggesting that these alleles reduce the likelihood of becoming a CHC. Those that were previously linked to increased Alzheimer’s Disease (AD) risk have much stronger effects on healthy longevity, indicating a broad association with healthy longevity beyond escaping AD. None of the tested disease-associated alleles were enriched in CHCs, possibly because AD-protective alleles were associated with major risk-increasing effects of other diseases, highlighting a complex network of interactions between disease and longevity associations in the *HLA* region.

## List of abbreviations

AD: Alzheimer’s Disease
AS: Ankylosing Spondylitis
ATD: Autoimmune Thyroid Disorders
CD: Celiac Disease
CER: Centenarian Effect Ratio
CHC: Cognitively Healthy Centenarians
CI: Confidence Interval
FDR: False Discovery Rate
GWAS: Genome-Wide Association Study
HLA: Human Leukocyte Antigen
LD: Linkage Disequilibrium
MS: Multiple Sclerosis
OR: Odds Ratio
PMR-GCA: Polymyalgia Rheumatica or Giant Cell Arteritis
RA: Rheumatoid Arthritis
SNP: Single Nucleotide Polymorphism
T1D: Type 1 Diabetes
UC: Ulcerative Colitis
UKB: UK Biobank

## Declarations

### Ethics approval and consent to participate

The medical ethics committee of the Amsterdam UMC approved all studies. All participants and/or their legal representatives provided written informed consent for participation in clinical and genetic studies.

### Consent for publication

Not applicable.

### Availability of data and materials

The dataset generated and analyzed during the current study are available in the Alzheimer’s Genetics Hub repository (https://alzheimergenetics.org/), on reasonable request.

## Funding

Research of the Alzheimer Center Amsterdam is part of the neurodegeneration research program of Amsterdam Neuroscience. Alzheimer Center Amsterdam is supported by Stichting Alzheimer Nederland and Stichting VUmc fonds.

DAS is supported by a VIDI Award (received by HH; WO/ZonMW # 09150172010083). NT, SvdL, MR, HH, and WvdF are recipients of ABOARD. ABOARD is a public–private partnership receiving funding from ZonMW National Dementia program (#73305095007) and Health∼Holland, Topsector Life Sciences & Health (PPP-allowance;#LSHM20106). More than 30 partners, including de Hersenstichting (Dutch Brain Foundation) participate in ABOARD (https://www.alzheimer-nederland.nl/onderzoek/projecten/aboard).

### Authors’ contributions

DAS analyzed and interpreted the data and was the major contributor in writing the manuscript. NT, MHul, AS, MR, SvdL and HH contributed substantially to the data analysis and interpretation and were contributors in writing the manuscript. NvdS, MHui, YP, WvdF, ES, and BU made substantial contributions to the conception of the work and acquisition of the data. All authors read and approved the final manuscript.

## Data Availability

The dataset generated and analyzed during the current study are available in the Alzheimers Genetics Hub repository (https://alzheimergenetics.org/), on reasonable request.

## Additional Figures and Tables

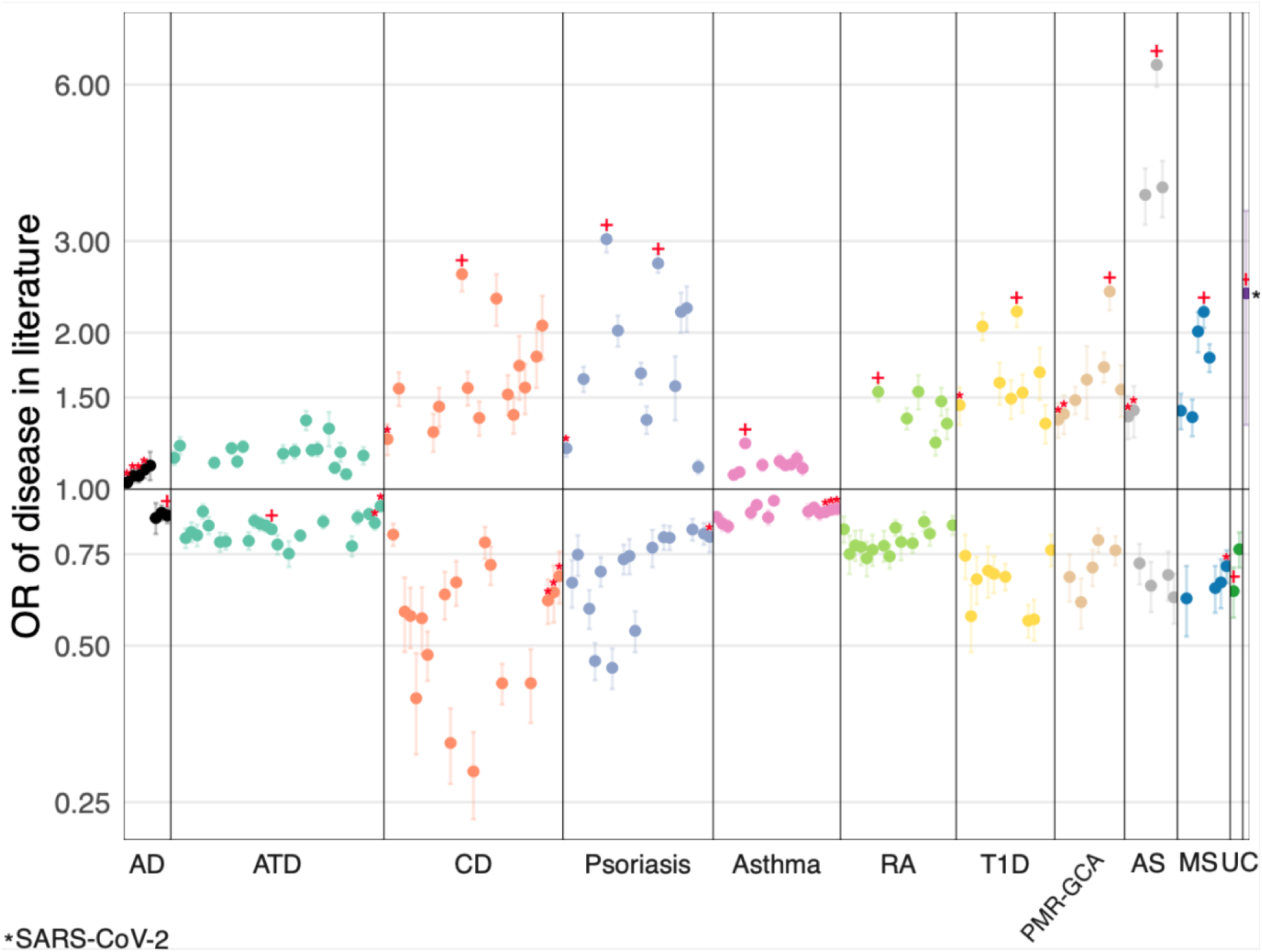

**Additional figure 1**. Odds Ratio with 95% CI of prior allele-disease associations in literature: AD-GWAS (12), UK Biobank (6) and SARS-CoV-2 (27), in a logarithmic scale. Red stars mark alleles that were significantly less frequent in CHCs compared to middle-aged healthy individuals. Red crosses indicate most-significant allele-disease associations per disease.

**Additional table 1 (additional_table1.csv).** Summary statistics for 59 unique HLA alleles, in 194 allele-disease associations. Allele name (Allele), allele frequency in study cohort (Freq.study), allele frequency in CHC (Freq. CHCs), allele frequency in middle-aged healthy individuals (Freq. Controls), Odds Ratio of becoming a CHC with 95% confidence intervals (ORs CHCs, OR_CHC_UpperCI, OR_CHC_LowerCI), p-value of association with becoming a CHC (p-value_CHC), false-discovery-rate corrected p-value of association (FDR_CHC), associated disease (Disease), Centenarian Effect Ratio with 95% CI (CER, CER_lowerCI, CER_upperCI), p-value of CER divergence from 0 (p-value_CER), FDR corrected p-value of p-value_CER (FDR_CER), allele frequency in literature study (Freq.literature), OR of disease association with 95% CI (OR_literature, OR_literature_LowerCI, OR_literature_UpperCI) and p-value of disease association in literature (p-value_literature).

## References

1. Kim ME, Lee JS. Immune Diseases Associated with Aging: Molecular Mechanisms and Treatment Strategies. Int J Mol Sci. 2023 Oct 25;24(21):15584.

2. Zheng Y, Liu Q, Goronzy JJ, Weyand CM. Immune aging – A mechanism in autoimmune disease. Semin Immunol. 2023 Sep;69:101814.

3. Timmers PR, Mounier N, Lall K, Fischer K, Ning Z, Feng X, et al. Genomics of 1 million parent lifespans implicates novel pathways and common diseases and distinguishes survival chances. eLife. 2019 Jan 15;8:e39856.

4. Melzer D, Pilling LC, Ferrucci L. The genetics of human ageing. Nat Rev Genet. 2020 Feb;21(2):88–101.

5. Horton R, Wilming L, Rand V, Lovering RC, Bruford EA, Khodiyar VK, et al. Gene map of the extended human MHC. Nat Rev Genet. 2004 Dec;5(12):889–99.

6. Butler-Laporte G, Farjoun J, Nakanishi T, Lu T, Abner E, Chen Y, et al. HLA allele-calling using multi-ancestry whole-exome sequencing from the UK Biobank identifies 129 novel associations in 11 autoimmune diseases. Commun Biol. 2023 Nov 3;6(1):1113.

7. Padyukov L. Genetics of rheumatoid arthritis. Semin Immunopathol. 2022 Jan;44(1):47– 62.

8. Mathew A, Bhagavaldas MC, Biswas R, Biswas L. Genetic risk factors in ankylosing spondylitis: Insights into etiology and disease pathogenesis. Int J Rheum Dis. 2024 Jan;27(1):e15023.

9. Wang QL, Wang TM, Deng CM, Zhang WL, He YQ, Xue WQ, et al. Association of HLA diversity with the risk of 25 cancers in the UK Biobank. eBioMedicine. 2023 Jun;92:104588.

10. Berryman MA, Ilonen J, Triplett EW, Ludvigsson J. Important denominator between autoimmune comorbidities: a review of class II HLA, autoimmune disease, and the gut. Front Immunol. 2023 Sep 26;14:1270488.

11. Hollenbach JA, Norman PJ, Creary LE, Damotte V, Montero-Martin G, Caillier S, et al. A specific amino acid motif of HLA-DRB1 mediates risk and interacts with smoking history in Parkinson’s disease. Proc Natl Acad Sci U S A. 2019 Apr 9;116(15):7419–24.

12. Bellenguez C, Küçükali F, Jansen IE, Kleineidam L, Moreno-Grau S, Amin N, et al. New insights into the genetic etiology of Alzheimer’s disease and related dementias. Nat Genet. 2022 Apr;54(4):412–36.

13. Le Guen Y, Luo G, Ambati A, Damotte V, Jansen I, Yu E, et al. Multiancestry analysis of the HLA locus in Alzheimer’s and Parkinson’s diseases uncovers a shared adaptive immune response mediated by HLA-DRB1*04 subtypes. Proc Natl Acad Sci U S A. 2023 Sep 5;120(36):e2302720120.

14. 14. International Multiple Sclerosis Genetics Consortium. Multiple sclerosis genomic map implicates peripheral immune cells and microglia in susceptibility. Science. 2019 Sep 27;365(6460):eaav7188.

15. Li X, Feng X, Sun X, Hou N, Han F, Liu Y. Global, regional, and national burden of Alzheimer’s disease and other dementias, 1990–2019. Front Aging Neurosci. 2022 Oct 10;14:937486.

16. Corrada MM, Brookmeyer R, Paganini-Hill A, Berlau D, Kawas CH. Dementia incidence continues to increase with age in the oldest old: the 90+ study. Ann Neurol. 2010 Jan;67(1):114–21.

17. Holstege H, Beker N, Dijkstra T, Pieterse K, Wemmenhove E, Schouten K, et al. The 100-plus Study of cognitively healthy centenarians: rationale, design and cohort description [Internet]. Neuroscience; 2018 [cited 2024 Dec 2]. Available from: http://biorxiv.org/lookup/doi/10.1101/295287

18. Tesi N, van der Lee S, Hulsman M, van Schoor NM, Huisman M, Pijnenburg Y, et al. Cognitively healthy centenarians are genetically protected against Alzheimer’s disease. Alzheimers Dement J Alzheimers Assoc. 2024 Jun;20(6):3864–75.

19. Barker DJ, Maccari G, Georgiou X, Cooper MA, Flicek P, Robinson J, et al. The IPD-IMGT/HLA Database. Nucleic Acids Res. 2023 Jan 6;51(D1):D1053–60.

20. Robinson J, Barker DJ, Marsh SGE. 25 years of the IPD-IMGT/HLA Database. HLA. 2024 Jun;103(6):e15549.

21. Trowsdale J, Knight JC. Major histocompatibility complex genomics and human disease. Annu Rev Genomics Hum Genet. 2013;14:301–23.

22. Marsh SGE, Albert ED, Bodmer WF, Bontrop RE, Dupont B, Erlich HA, et al. Nomenclature for factors of the HLA system, 2010. Tissue Antigens. 2010 Apr;75(4):291–455.

23. Kennedy AE, Ozbek U, Dorak MT. What has GWAS done for HLA and disease associations? Int J Immunogenet. 2017 Oct;44(5):195–211.

24. Naito T, Okada Y. HLA imputation and its application to genetic and molecular fine-mapping of the MHC region in autoimmune diseases. Semin Immunopathol. 2022 Jan;44(1):15–28.

25. Lio D, Pes GM, Carru C, Listì F, Ferlazzo V, Candore G, et al. Association between the HLA-DR alleles and longevity: a study in Sardinian population. Exp Gerontol. 2003 Mar;38(3):313–7.

26. Joshi PK, Pirastu N, Kentistou KA, Fischer K, Hofer E, Schraut KE, et al. Genome-wide meta-analysis associates HLA-DQA1/DRB1 and LPA and lifestyle factors with human longevity. Nat Commun. 2017 Oct 13;8(1):910.

27. Augusto DG, Murdolo LD, Chatzileontiadou DSM, Sabatino JJ, Yusufali T, Peyser ND, et al. A common allele of HLA is associated with asymptomatic SARS-CoV-2 infection. Nature. 2023 Aug 3;620(7972):128–36.

28. Hoogendijk EO, Deeg DJH, Poppelaars J, Van Der Horst M, Broese Van Groenou MI, Comijs HC, et al. The Longitudinal Aging Study Amsterdam: cohort update 2016 and major findings. Eur J Epidemiol. 2016 Sep;31(9):927–45.

29. Van Der Flier WM, Scheltens P. Amsterdam Dementia Cohort: Performing Research to Optimize Care. Perry G, Avila J, Tabaton M, Zhu X, editors. J Alzheimers Dis. 2018 Mar 13;62(3):1091–111.

30. Rademaker MC, De Lange GM, Palmen SJMC. The Netherlands Brain Bank for Psychiatry. In: Handbook of Clinical Neurology [Internet]. Elsevier; 2018 [cited 2024 Dec 2]. p. 3–16. Available from: https://linkinghub.elsevier.com/retrieve/pii/B9780444636393000013

31. Willemsen G, De Geus EJC, Bartels M, Van Beijsterveldt CEMT, Brooks AI, Estourgie-van Burk GF, et al. The Netherlands Twin Register Biobank: A Resource for Genetic Epidemiological Studies. Twin Res Hum Genet. 2010 Jun 1;13(3):231–45.

32. Loonstra FC, De Ruiter LRJ, Doesburg D, Lam KH, Van Lierop ZYGJ, Moraal B, et al. Project Y: The search for clues explaining phenotype variability in MS. Mult Scler Relat Disord. 2022 Jan;57:103337.

33. Das S, Forer L, Schönherr S, Sidore C, Locke AE, Kwong A, et al. Next-generation genotype imputation service and methods. Nat Genet. 2016 Oct;48(10):1284–7.

34. Anderson CA, Pettersson FH, Clarke GM, Cardon LR, Morris AP, Zondervan KT. Data quality control in genetic case-control association studies. Nat Protoc. 2010 Sep;5(9):1564–73.

35. The 1000 Genomes Project Consortium, Corresponding authors, Auton A, Abecasis GR, Steering committee, Altshuler DM, et al. A global reference for human genetic variation. Nature. 2015 Oct 1;526(7571):68–74.

36. Zheng X, Shen J, Cox C, Wakefield JC, Ehm MG, Nelson MR, et al. HIBAG--HLA genotype imputation with attribute bagging. Pharmacogenomics J. 2014 Apr;14(2):192– 200.

37. Price AL, Patterson NJ, Plenge RM, Weinblatt ME, Shadick NA, Reich D. Principal components analysis corrects for stratification in genome-wide association studies. Nat Genet. 2006 Aug;38(8):904–9.

38. Price AL, Zaitlen NA, Reich D, Patterson N. New approaches to population stratification in genome-wide association studies. Nat Rev Genet. 2010 Jul;11(7):459–63.

39. Benjamini Y, Hochberg Y. Controlling the False Discovery Rate: A Practical and Powerful Approach to Multiple Testing. J R Stat Soc Ser B Stat Methodol. 1995 Jan 1;57(1):289–300.

40. Purcell S, Neale B, Todd-Brown K, Thomas L, Ferreira MAR, Bender D, et al. PLINK: a tool set for whole-genome association and population-based linkage analyses. Am J Hum Genet. 2007 Sep;81(3):559–75.

41. Tesi N, Van Der Lee SJ, Hulsman M, Jansen IE, Stringa N, Van Schoor N, et al. Centenarian controls increase variant effect sizes by an average twofold in an extreme case–extreme control analysis of Alzheimer’s disease. Eur J Hum Genet. 2019 Feb;27(2):244–53.

42. Matteson EL, Dejaco C. Polymyalgia Rheumatica. Ann Intern Med. 2017 May 2;166(9):ITC65–80.

43. Tan DJ, Walters EH, Perret JL, Lodge CJ, Lowe AJ, Matheson MC, et al. Age-of-asthma onset as a determinant of different asthma phenotypes in adults: a systematic review and meta-analysis of the literature. Expert Rev Respir Med. 2015 Jan 2;9(1):109–23.

44. Calcaterra V, Nappi RE, Regalbuto C, De Silvestri A, Incardona A, Amariti R, et al. Gender Differences at the Onset of Autoimmune Thyroid Diseases in Children and Adolescents. Front Endocrinol. 2020 Apr 17;11:229.

45. Prosperini L, Lucchini M, Ruggieri S, Tortorella C, Haggiag S, Mirabella M, et al. Shift of multiple sclerosis onset towards older age. J Neurol Neurosurg Psychiatry. 2022 Oct;93(10):1137–9.

46. Corrada MM, Hayden KM, Paganini-Hill A, Bullain SS, DeMoss J, Aguirre C, et al. Age of onset of hypertension and risk of dementia in the oldest-old: The 90+ Study. Alzheimers Dement. 2017 Feb;13(2):103–10.

47. Redondo MJ, Concannon P. Genetics of Type 1 Diabetes Comes of Age. Diabetes Care. 2020 Jan 1;43(1):16–8.

48. Brophy S, Calin A. Ankylosing spondylitis: interaction between genes, joints, age at onset, and disease expression. J Rheumatol. 2001 Oct;28(10):2283–8.

49. Caio G, Volta U, Sapone A, Leffler DA, De Giorgio R, Catassi C, et al. Celiac disease: a comprehensive current review. BMC Med. 2019 Dec;17(1):142.

50. Queiro R, Tejón P, Alonso S, Coto P. Age at disease onset: a key factor for understanding psoriatic disease. Rheumatology. 2014 Jul;53(7):1178–85.

